# Influence of parental anthropometry and gestational weight gain on intrauterine growth and neonatal outcomes: Findings from the MAI cohort study in rural India

**DOI:** 10.1101/2023.04.06.23288237

**Authors:** Mugdha Deshpande, Demi Miriam, Nikhil Shah, Neha Kajale, Jyotsna Angom, Jasmin Bhawra, Ketan Gondhalekar, Anuradha Khadilkar, Tarun Katapally

## Abstract

**Background:** Poor foetal growth and subsequent low birth weight are associated with an increased risk for disease later in life. Identifying parental factors that determine foetal growth are important to curbing intergenerational malnutrition, especially among disadvantaged populations in the global south where undernutrition rates are high. The objective of this study was to assess the relationships between parental biometry, intrauterine growth and neonatal outcomes, while factoring in socioeconomic status of historically disadvantaged households in rural India

**Material and Methods:** Using data from the prospective longitudinal cohort, pregnant women from rural Pune, India (n = 134) were assessed between August 2020 and November 2022. Data on socio-demography, ultrasound measurements, parental and foetal anthropometry were collected. Multiple linear regression models were run to predict determinants of foetal intrauterine and neonatal growth (*p* value<0.05). The dependent variables were ultrasound measurements and neonatal biometry, and independent variables were gestational weight gain, parental and mid-parental height.

**Results:** Mean(±SD) maternal age, maternal height, paternal height and mid-parental height were 22.8±3.7 years, 153.6±5.5cm, 165.9±6.5cm and 159.1±8.7cm, respectively. Pre-pregnancy body mass index and gestational weight gain was 20.5±4.0 kg/m^2^ and 9.8±3.7kg respectively. Mid-parental height and gestational weight gain were strongly correlated with neonatal growth and foetal intrauterine growth (*p*<0.05); however, the correlation peaked at 28 weeks of gestation(*p*<0.05). Gestational weight gain (B=28.7, *p*=0.00) and mid-parental height (B=14.3, *p*=0.00) were identified as strong determinants of foetal-intrauterine growth and neonatal anthropometry at birth. Maternal height was found to influence length of male neonate (B=0.18, *p*=0.00), whereas, paternal height influenced length of the female neonate (B=0.11, *p*=0.01).

**Conclusion:** Parental socio-economic status, biometry and maternal gestational weight gain influence growth of the child starting from the intrauterine period. Our study underlines the need for interventions during pre-pregnancy, as well as during pregnancy, for optimal weight gain and improved foetal and neonatal outcomes.

## Introduction

Poor foetal growth and subsequent low birth weight are associated with an increased risk for disease later in life(1). Foetal development is a reflection of the prenatal environment, which constitutes shared maternal and paternal genetic, environmental and nutritional factors(2). It has been inferred from past studies that environmental and genetic factors influence neonatal outcomes at birth(3). Nutritional and socioeconomic status of parents, and maternal age are amongst the few factors that impact foetal and neonatal growth(4). This implies that physiological adaptations brought on by genetic exposures during the prenatal stage affect the long-term growth and health of a child(5). Therefore, identifying the factors impacting foetal growth will help to understand factors affecting neonatal size and survival(6).

Maternal anthropometry (height, pre-pregnancy weight, and gestational weight gain (GWG)) has a strong impact on foetal growth as well as birth weight and body composition of the neonate(7,8). It has been documented that maternal pre-pregnancy weight and gestational weight gain are positively correlated with neonatal size(7,8). Pre-pregnancy body mass index (BMI) represents chronic maternal nutritional status; however, gestational weight gain reflects both acute nutritional status and the growth of tissues(9).

Paternal anthropometry influences foetal-neonatal growth; however, studies on the impact of father’s height on the longitudinal pattern of foetal growth are lacking. Research shows that parental heights affect the growth of their offspring(10) which was first described by Galton (11) as mid-parental height (MPH).However, studies assessing the relationship of MPH with foetal and neonatal growth are scarce.

To differentiate between foetuses that are at the extreme ends of the growth spectrum (e.g., small for gestational age vs. large for gestational age), MPH along with individual anthropometric characteristics of the mother and father could be studied together with foetal growth parameters such as: crown-rump length, femur length, biparietal diameter, head circumference, abdominal circumference and estimated foetal weight. These measures reflect foetal growth(12) as evidenced in ultrasound measurements and neonatal growth parameters by neonatal anthropometry including neonatal birth weight, recumbent length, chest circumference, mid upper arm circumference and head circumference at birth(4).

From a public health perspective, identifying factors which impact foetal and neonatal growth is particularly crucial in historically disadvantaged households in rural India-an low-middle income setting, where 12.0% to 78.4% of neonates are born small for their gestational age(13). In India, 25% of women in the reproductive age in rural areas are underweight (National Family Health Survey-5, India (2019-20)), thus, identifying parental factors that determine foetal growth are important in order to curb intergenerational malnutrition. This longitudinal study aimed to assess the relationship between parental biometry, intrauterine growth and neonatal outcomes, as well as the determinants of foetal and neonatal growth among a rural Indian population of pregnant mothers near Pune, India.

## Methods

### Study design and subjects

This study was a part of the Mother and Infant cohort (MAI: Mother in Marathi-a native language of Maharashtra), a single-centre, prospective, observational, longitudinal community-based cohort study conducted in rural areas near Pune city in Maharashtra, India.

Sample size calculation: A-priori sample size was calculated to 175 after accounting for 20% attrition rate, 0.8 power with an effect size f^2^ of 0.1 and alpha 0.05 using linear multiple regression fixed model (F test family) G power software (Version 3.1.9.7). After enrolling 175 participants, complete data on 134 was obtained for which post-hoc power was re-calculated (proportions: difference from constant (G power software), effect size f^2^ of 0.1 and alpha 0.05). The power of the study remained unchanged at 0.8 with 134 final samples.

Based on the study inclusion criteria, healthy pregnant women attending antenatal health camps in primary healthcare centres and Anganwadi centres (i.e., rural childcare centres) in their first trimester, who planned to deliver within a 150km radius (to ensure the feasibility of post-delivery measurement) were enrolled after obtaining consent. Women with comorbidities such as diabetes mellitus, and hypertension, with a gestational age of more than 12 weeks, those bearing twins, women who planned to deliver beyond pre-decided distance limits, and those who did not consent were excluded from the study.

A total of 553 pregnant women were screened, after which 175 participants were being enrolled. After accounting for drop-outs, abortions, still-births, and intrauterine deaths, 41 records were excluded. Thus, the final study included 134 triads (mother, father, and child). Details of recruitment are explained in figure 1.

**Figure 1:**
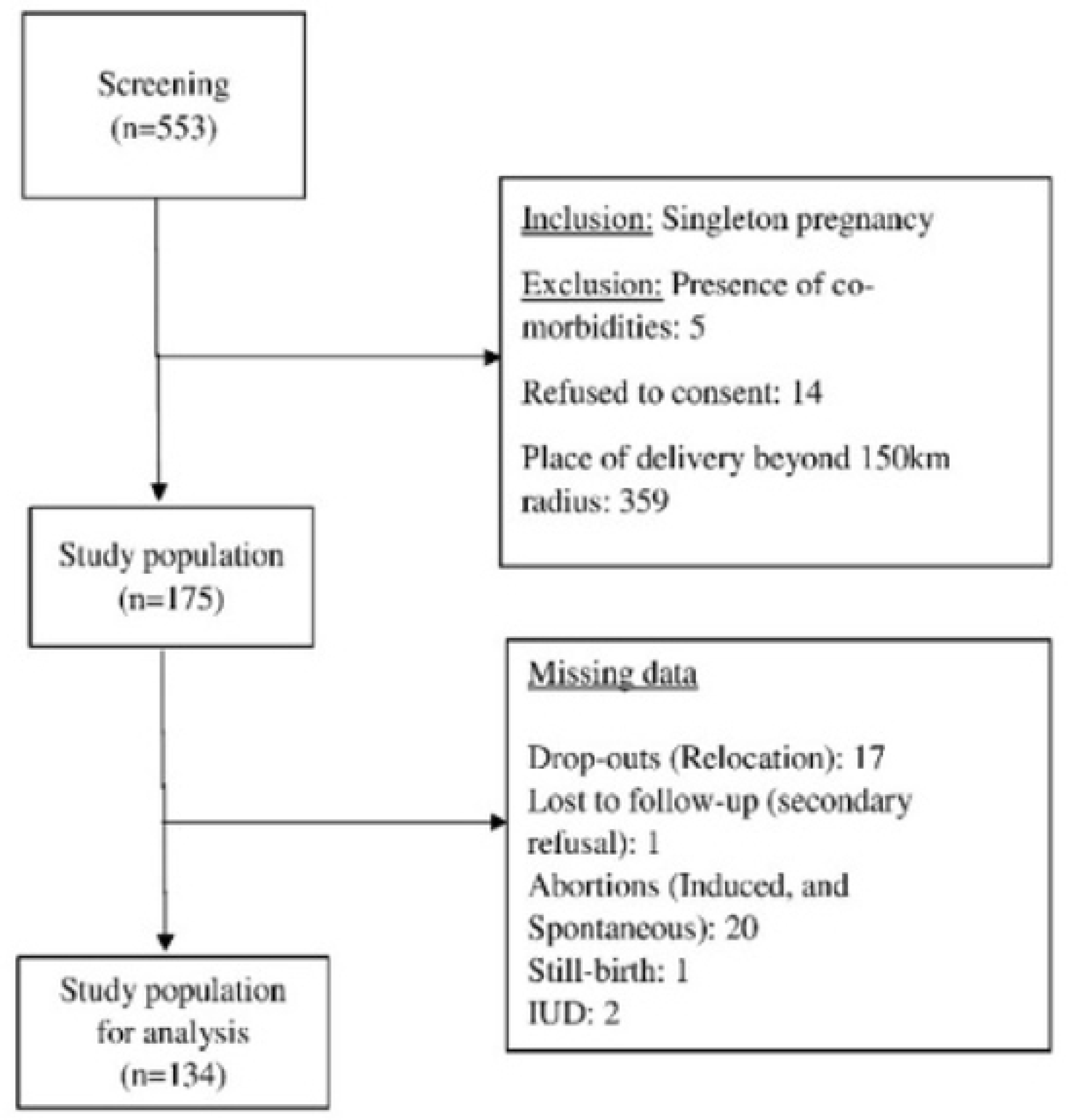
Recruitment flowchart for MAI cohort.

### Data collection

Data collection was performed between August 2020 and November 2022. All participants gave written informed consent before any study procedures were performed. Ethics approval was obtained from the Institutional ethics committee (EC registration no.: ECR/352/Inst/MH/2013/RR-19). Data were collected at five time points during clinic visits (Visit (V)1-V5): V1 - first trimester (8-12 weeks Gestational Age (GA)), V2 - second trimester (18-22 weeks GA), V3 - third trimester (28-32 weeks GA), V4 - before delivery and post-delivery (within 24 hours to 10 days). Follow-up was conducted via phone calls and messages every month until 34 weeks of gestation, and then once a week from 34 weeks until after delivery to ensure study compliance.

### 2.1 Parental data

Family socioeconomic status was recorded using the New Socioeconomic Classification (SEC) system according to socioeconomic classification 2011(14). The New SEC system scores families based on the number of owned material possessions, which include consumer assets ranging from electricity connection to owning agricultural land and education of the chief earner of the family(14). The SEC scores were recoded as a binary variable in statistical analysis for multivariate models as high and low socioeconomic status. Maternal and paternal height was recorded to the nearest 0.5cm using a SECA stadiometer. Maternal weight was recorded using an electronic weighing scale to the nearest 0.5kg. For calculating maternal pre-pregnancy BMI, weight was self-reported by participants. The World Health Organization guidelines for BMI were used, where underweight status was defined as BMI less than 18.5kg/m^2^, normal weight status as 18.5-24.9 kg/m^2^, and overweight and obese statuses were between 25-29.9 kg/m^2^ and >30 kg/m^2^ respectively(15).

The Institute of Medicine (IOM) provided guidelines for GWG based on pre-pregnancy BMI in 2009(15). A weight gain of 12.5-18kg is recommended for underweight women, 11.5-16kg for normal weight status women, and 7-11.5kg and 5-9 kg for, overweight and obese BMI category women, respectively(15).

BMI was computed using the formula:

*BMI (kg/m^2^) = weight (kg)/ height (m)^2^*

GWG was computed by the formula:

*GWG (kg)= Weight at V4 (kg)-Weight at V1(kg)*

V1: weight at first-trimester visit and V4: Weight just before delivery

Mid-parental height was computed using the formula:

*(Paternal height (cm)+ maternal height (cm)/2) + 6.5 for boys and;*

*(Paternal height (cm)+ maternal height (cm)/2) - 6.5 for girls*

### 2.2 Foetal data

Foetal ultrasound examinations were conducted at four time points during the course of pregnancy: once in early (GA:8 ± 2 weeks) and mid-pregnancy (GA:20 ± 2 weeks) and twice in late pregnancy (GA: 30 ± 2 weeks and 36 ± 2 weeks). The first ultrasound examination was not included in the final analysis since it did not include growth characteristics. Foetal growth parameters of interest were head circumference, abdominal circumference, bi-parietal diameter, femur length in centimetres and estimated foetal weight in grams which were uniformly reported at GA of 18 weeks and onwards. Bi-parietal diameter was measured in an axial view with foetal positioning in direct occiput transverse. Measurement was taken from the inner margin of the opposite skull table to the outer surface of the skull table. Head circumference was measured along the outer perimeter of the calvarium at the same level as that of bi-parietal diameter. Femur length measurement was made along the axis of diaphysis from later epicondyle to the tip of greater trochanter in a straight line. Abdominal circumference was measured at the level of the foetal liver, taking into view the umbilical part of the left portal vein in the liver. Measurement was made from the outer edge of one side of the foetal abdomen to the other side edge(16).

Estimated foetal weight was calculated using Hadlock’s formula:

*(log10 estimated foetal weight = 1.5662) 0.0108 (Head circumference) + 0.0468 (Abdominal circumference) + 0.171 (Femur length) + 0.00034 (Head circumference)2) 0.003685 (Abdominal circumference*Femur length))(17)*

### 2.3 Neonatal data

Neonatal anthropometry as per world health organization protocol(18) and delivery details (e.g. Type of delivery, membrane rupture, APGAR score, etc.) were collected within 10 days of delivery by researchers who were trained in measuring anthropometry. All the equipment was calibrated regularly using industry standards(19). Neonatal birth weight was recorded using an electronic weighing scale that had tarring capacity, and the weight was measured with a high precision (within 0.1kg) by placing the baby on the weighing scale and after adjusting for neonate’s minimal clothing.

Recumbent length was recorded using the SECA infantometer, where the neonate’s diapers were removed since it made it difficult to hold the neonate’s legs together. Two researchers sat on either side of the infantometer, where one held the baby’s feet together and the other adjusted the head of the neonate and moved the headboard. The position of the neonate was such that the crown touched the headboard and the distance between the ear canal to lower border of eye socket formed a vertical line which was perpendicular to horizontal board (vertical Frankfurt plane). After adjusting for neonate’s legs and hips, gentle pressure was applied to straighten the neonate’s knees and the footboard was positioned against soles of the feet and toes pointing outward to measure length to the nearest 0.1cm.

For measuring head circumference, a non-bendable, non-stretchable measuring tape was used to cover the fullest protuberance of skull and eyebrows after carefully positing the tape sideways and then the tape was snugly pulled to compress hair and skin and the reading was recorded to the nearest 0.1cm. For measuring the mid-upper arm circumference, a mid-point was placed between the acromion and the olecranon process of the elbow by bending the neonate’s forearm to 90° at elbow with palm facing up so that olecranon stood out from the elbow. Then circumference was measured by relaxing the neonate’s elbow in an extended position, carefully observing that the muscles didn’t tighten, by snugly measuring the mid-point using a non-bendable, non-stretchable SECA measuring tape to the nearest 0.1cm. Chest Circumference was recorded after removing neonate’s top clothing and marking a horizontal line at xiphisternum where ribs meet sternum and placing a non-bendable, non-stretchable SECA measuring tape such that the upper part of tape touched the marking to measure chest circumference to the nearest 0.1cm. Small for gestational age was defined as birth length and birth weight of less than 10^th^ percentile of the average weight and length for gestational age in neonates of same gender(20). Low birth weight was considered to be birth weight of less than 2.5kg(21). Preterm birth was defined as birth of the foetus before 37 weeks of gestation(22).

### Statistical analysis

Statistical analysis was performed using Statistical Package for Social Sciences version 25. Continuous variables were expressed as mean and standard deviation and categorical variables were expressed in percentages. All outcome variables were tested for normality before performing analysis. T-Tests were performed to assess differences in means of parental anthropometry and foetal-neonatal biometry. Test of proportions was used to assess significance in differences between two groups. Median values of parental anthropometry were considered since the participants who were under -2SD ranged from 3-4.5% which was inadequate for them to be compared to their counterparts. Linear association of variables was tested using Pearson’s correlation for normal data and Spearman’s correlation for non-normal data. Multiple linear regression models were run to predict determinants of foetal intrauterine growth and neonatal growth. The dependent variables were ultrasound measurements i.e., growth characteristics (femur length and estimated foetal weight) and neonatal biometry (birth weight and recumbent length) and the primary independent variables were GWG, MPH, maternal and paternal height. *p* value of <0.05 was considered statistically significant.

## Results

The overall sample included 134 triads of mothers, fathers and children (44% (59) male and 56% (75) female). Maternal age at marriage and at pregnancy was 20.2±3.1 years and 23.4±3.7 years, respectively, with minimum and maximum maternal age being 16 years and 35.4 years, respectively. In terms of education, 4.5% mothers had no official literacy (i.e., never attended school),65.6% obtained 10^th^/12^th^ grade qualifications, and 29.6% reported education of higher than 12th grade. In terms of employment, 16.4% mothers reported that they were employed until 28 weeks of gestation, while 83.6% mothers were home-makers (i.e., reported no formal employment). One-third of mothers (32.1%) belonged to families of low socio-economic status.

Based on pre-pregnancy BMI, it was observed that 36.6% of mothers were underweight (BMI<18.5kg/m^2^), 50.7% were of normal weight (18.5-24.9 kg/m^2^), 9% were classified with overweight status (25-29.9 kg/m^2^), and 3.7% were categorized to obese weight status (>30 kg/m^2^). Overall, 59.7% of mothers had inadequate GWG, with 33.6% gaining appropriate weight, and 6.7% gaining excess GWG, according to IOM guidelines. Of mothers who belonged to the underweight pre-pregnancy BMI category, 61.2% did not gain adequate weight during pregnancy as is shown in figure 2.

**Figure 2:**
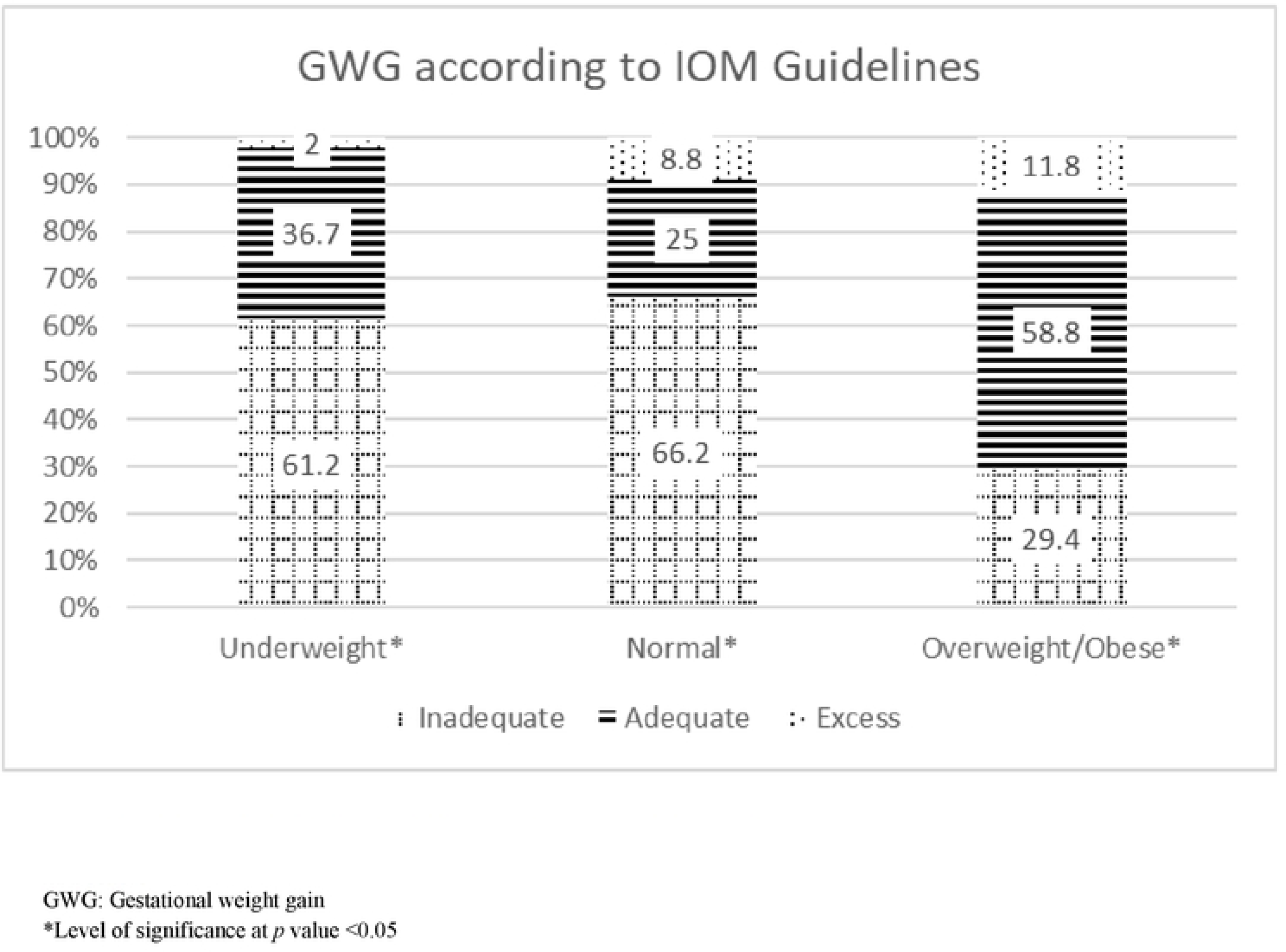
GWG according to IOM Guidelines in rural Indian pregnant women from MAI cohort.

In terms of numbers of historical pregnancies per woman, approximately 30% were nulliparous (first pregnancy), and almost 70% were primiparous (had previously given birth at least once). Of the total deliveries recorded within the sample, 20.1% were pre-term (deliveries of gestational age<37 weeks). The prevalence of small-for-gestational age computed at birth was found to be 42.5%, with a higher percentage noted among male neonates. Prevalence of low birth weight was found to be 23.9% with a larger proportion of female neonates weighing below 2.5kg. Maternal, paternal, and neonatal characteristics of the study population are summarized in table 2.

**Table 1:** Data collected at different time periods from MAI cohort.

2.1 Parental data
| Time frame | Data collected |
| --- | --- |
| I trimester (8-12 weeks) | Parental socio-economic data, maternal and paternal height, maternal weight at baseline, data from ultrasonography (USG) viability scan) |
| II trimester (18-22 weeks) | Maternal weight, data from USG (nuchal translucency scan to rule out genetic abnormalities and anomaly scan: growth parameters) |
| III trimester (28-32 weeks) | Maternal weight, data from USG (growth scan at 28 and 36 weeks of gestation: growth parameters) |
| Before delivery | Weight |
| Post-delivery (within 24 hours to 10 days) | Maternal and neonatal outcomes (birth weight, recumbent length, mid-upper arm circumference, head circumference and chest circumference) |
| **Table 1: Data collected at different time periods from MAI cohort** |  |

**Table 2:** Parental and neonatal characteristics of study population from MAI cohort

| Parameter | Mean±S.D. |
| --- | --- |
| Maternal age (years) | 23.4±3.7 |
| Maternal height (cm) | 153.6±5.6 |
| Paternal height (cm) | 166.0±6.4 |
| Mid-parental height (cm) | 159.1±8.7 |
| Maternal body mass index (kg/m<sup>2</sup>) | 20.5±4.0 |
| Gestational age (weeks) | 38.3±1.8 |
| Gestational weight gain (kg) | 9.9±3.7 |
| Neonatal birth weight (kg) | 2.6±0.4 |
| Neonatal Recumbent length (cm) | 48.7±2.6 |
| Neonatal Head circumference (cm) | 33.3±3.4 |
| Neonatal Chest circumference (cm) | 31.8±3.3 |
| Neonatal Mid-upper arm circumference (cm) | 9.3±1.1 |

Foetal growth parameters as assessed through foetal ultrasonography over the course of gestation at 20 weeks, 28 weeks and 36 weeks respectively are described in table 3. Foetal growth parameters showed a significant increase over the duration of pregnancy (p<0.05).

**Table 3:**
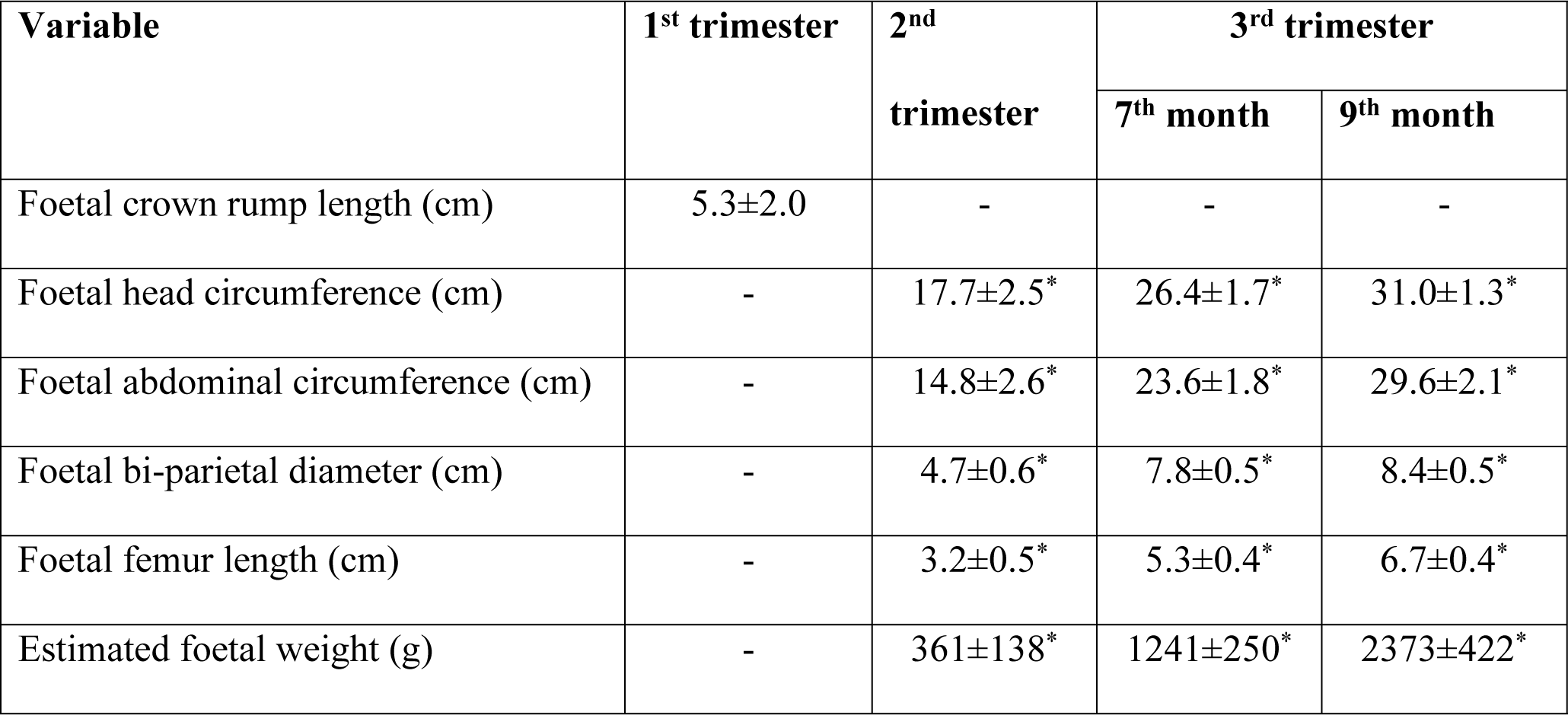
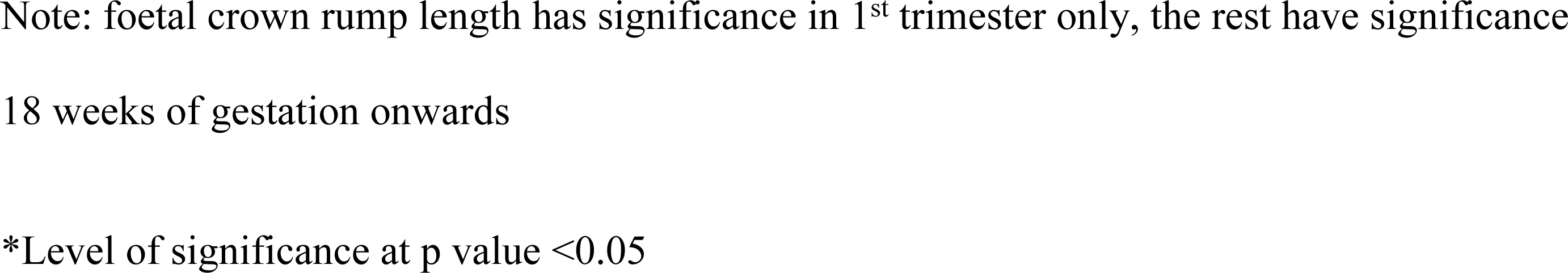
Foetal growth parameters in rural Indian pregnancies from MAI cohort

| Variable | 1<sup>st</sup> trimester | 2<sup>nd</sup> trimester | 3<sup>rd</sup> trimester |  |
| --- | --- | --- | --- | --- |
|  |  |  | 7<sup>th</sup> month | 9<sup>th</sup> month |
| Foetal crown rump length (cm) | 5.3±2.0 | - | - | - |
| Foetal head circumference (cm) | - | 17.7±2.5* | 26.4±1.7* | 31.0±1.3* |
| Foetal abdominal circumference (cm) | - | 14.8±2.6* | 23.6±1.8* | 29.6±2.1* |
| Foetal bi-parietal diameter (cm) | - | 4.7±0.6* | 7.8±0.5* | 8.4±0.5* |
| Foetal femur length (cm) | - | 3.2±0.5* | 5.3±0.4* | 6.7±0.4* |
| Estimated foetal weight (g) | - | 361±138* | 1241±250* | 2373±422* |

Note: foetal crown rump length has significance in 1<sup>st</sup> trimester only, the rest have significance 18 weeks of gestation onwards
\*Level of significance at p value <0.05

Foetal and neonatal growth was significantly different between mothers who gained adequate versus inadequate weight, with an exception of no difference observed between the two groups with respect to femur length, abdominal circumference, and neonatal head circumference (table 4). A large number of significant differences were seen between medians of paternal height as compared to the medians of maternal height (p < 0.05) as demonstrated in table 4. On comparison of medians of MPH, significant foetal and neonatal growth differences were also observed (p < 0.05).

**Table 4:**
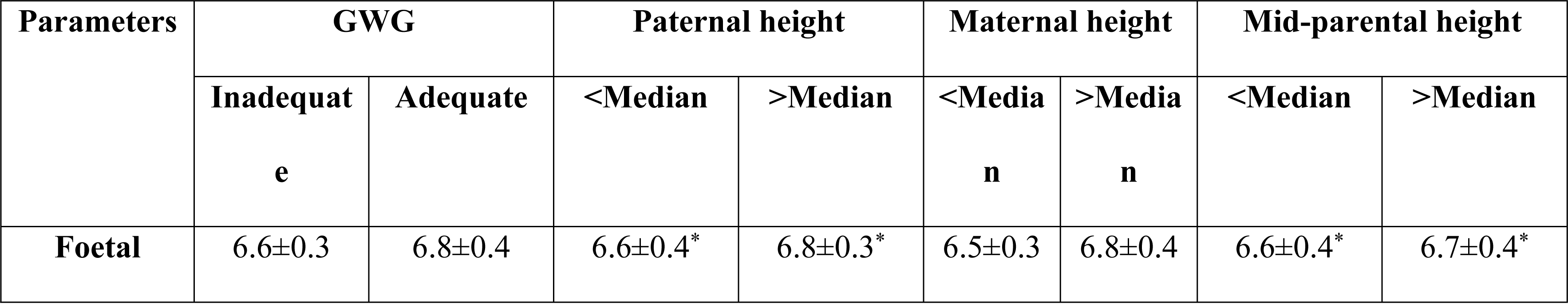
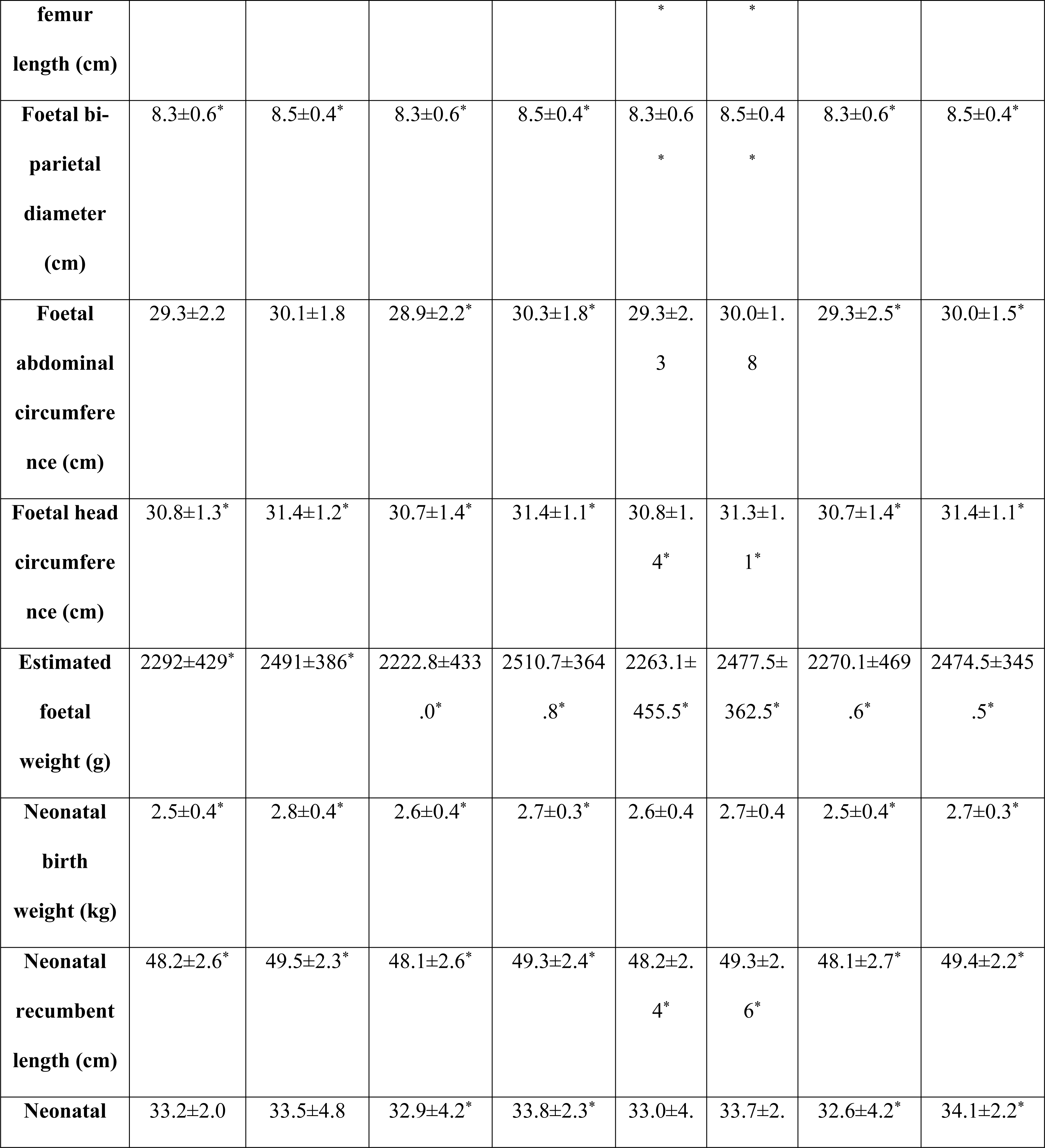
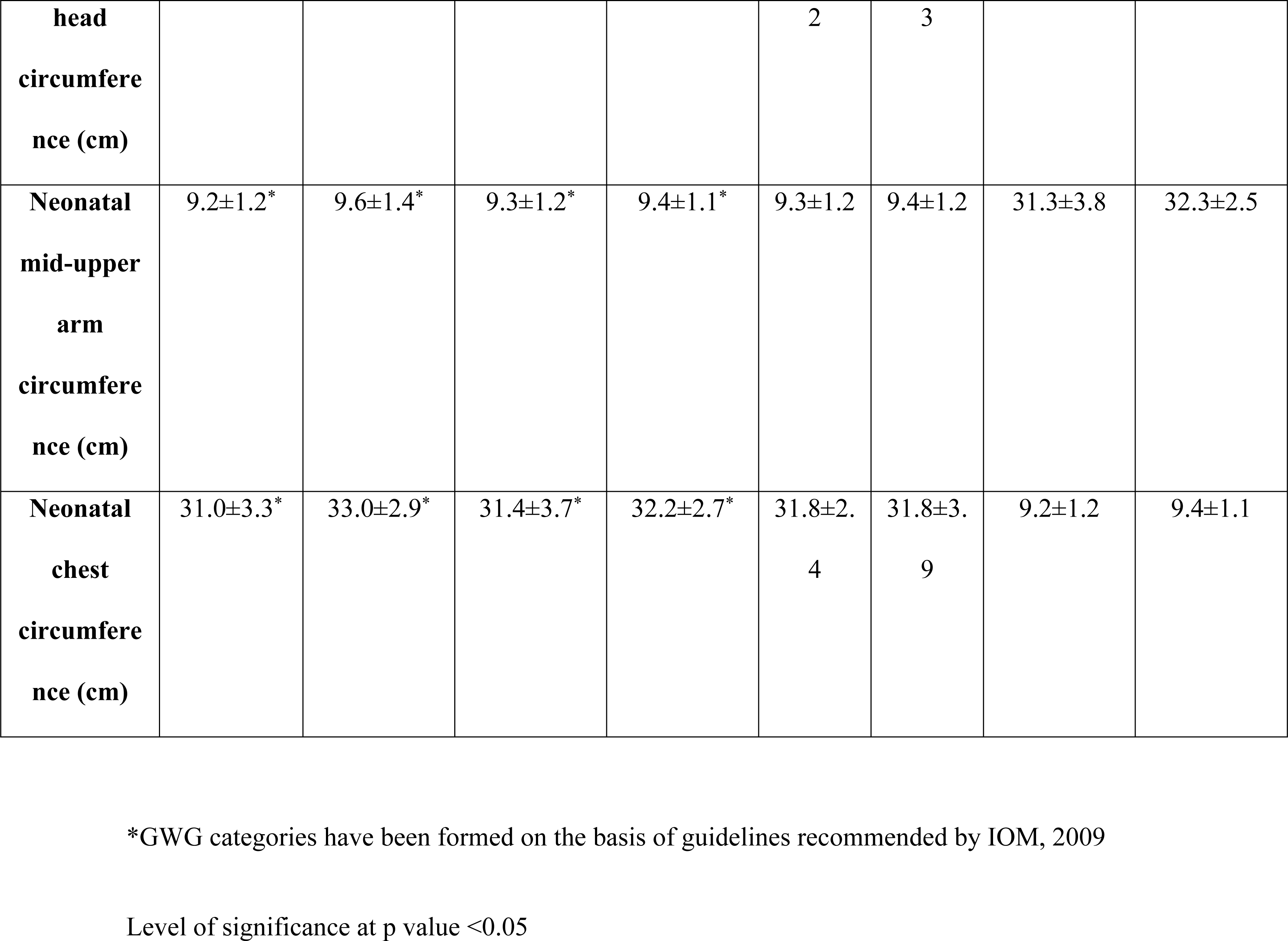
Difference in means of parental parameters and foetal-neonatal growth characteristics from MAI cohort

Significant correlations were observed between parental anthropometry and foetal-neonatal growth (Table 5); however, the correlation was significant only after 28 weeks of gestation with strongest correlation observed at 36 weeks of gestation. Maternal pre-pregnancy BMI significantly correlated with only femur length at 20 weeks of gestation and birth weight (p < 0.05). The relationship between the foetal and neonatal growth parameters with maternal height was significant only from 36 weeks of gestation (p< 0.05). Similarly, foetal and neonatal growth parameters were significantly correlated with paternal height from 36 weeks of gestation (p < 0.05).

**Table 5:**
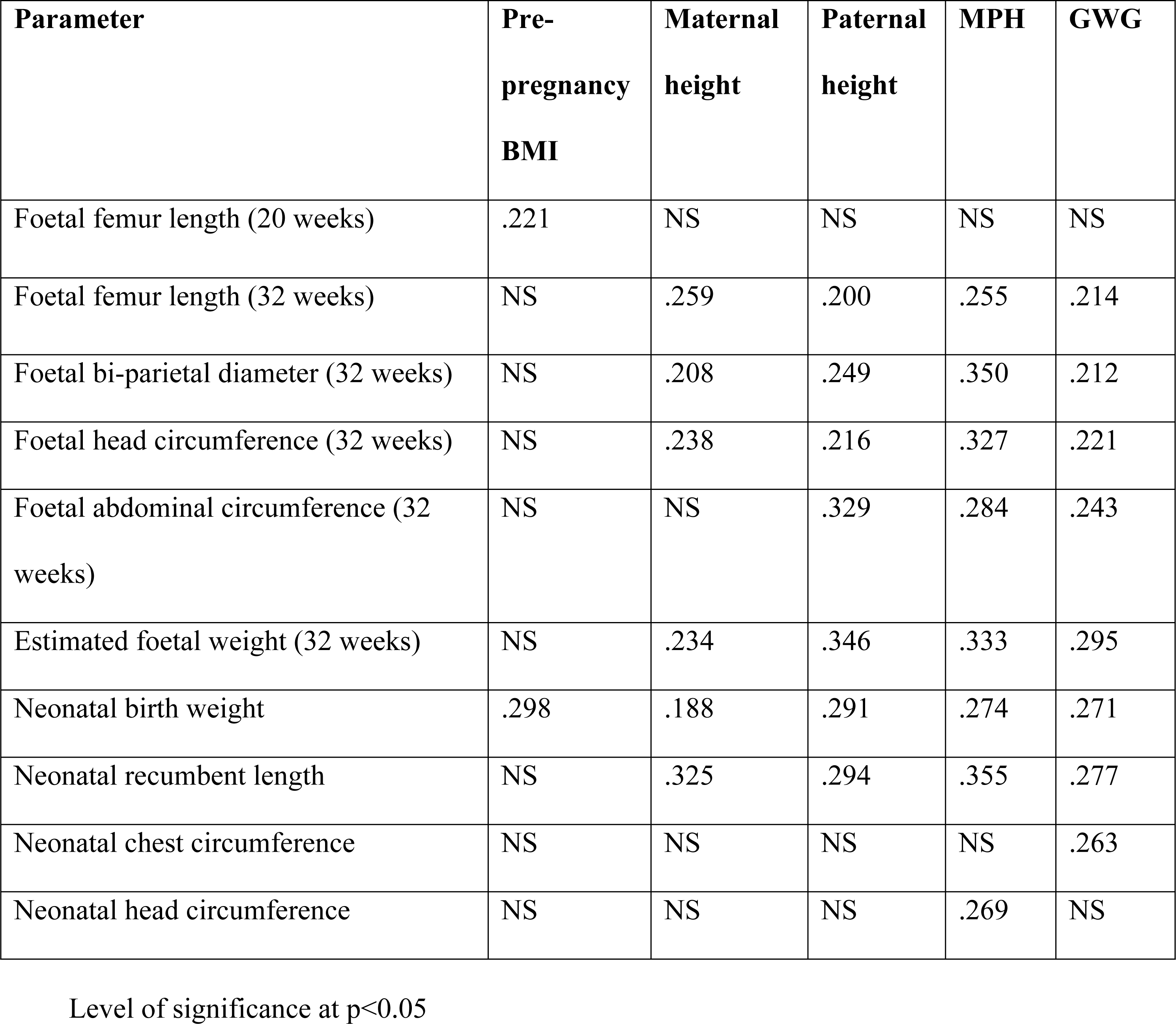
Correlation between parental and foetal-neonatal growth parameters from MAI cohort

| Parameter | Pre-pregnancy BMI | Maternal height | Paternal height | MPH | GWG |
| --- | --- | --- | --- | --- | --- |
| Foetal femur length (20 weeks) | .221 | NS | NS | NS | NS |
| Foetal femur length (32 weeks) | NS | .259 | .200 | .255 | .214 |
| Foetal bi-parietal diameter (32 weeks) | NS | .208 | .249 | .350 | .212 |
| Foetal head circumference (32 weeks) | NS | .238 | .216 | .327 | .221 |
| Foetal abdominal circumference (32 weeks) | NS | NS | .329 | .284 | .243 |
| Estimated foetal weight (32 weeks) | NS | .234 | .346 | .333 | .295 |
| Neonatal birth weight | .298 | .188 | .291 | .274 | .271 |
| Neonatal recumbent length | NS | .325 | .294 | .355 | .277 |
| Neonatal chest circumference | NS | NS | NS | NS | .263 |
| Neonatal head circumference | NS | NS | NS | .269 | NS |

Level of significance at $p < 0.05$

Multiple linear regression models were run to determine foetal-neonatal growth after adjusting for socioeconomic status (table 6). Socioeconomic status was added in layer one to observe the independent effect of low family socioeconomic status on the dependent variable with the control as high family socioeconomic status and again in layer two to observe its additional impact on the dependent variable in combination with other independent variables. GWG and MPH were significant predictors of foetal length and weight (p < 0.05). With every 1cm increase in paternal height, foetal weight increased by 19g. Similarly, with every 1 kg increase in GWG, foetal weight increased by 21g. Neonatal birth weight was significantly associated with GWG and MPH, whereas, neonatal recumbent length was significantly associated with parental anthropometry overall (p < 0.05). On further analysis, paternal height was significantly associated with recumbent length of female neonates (B: 0.110, p < 0.05) whereas, the recumbent length of male neonates was significantly determined by maternal height (B: 0.151, p < 0.05).

**Table 6:**
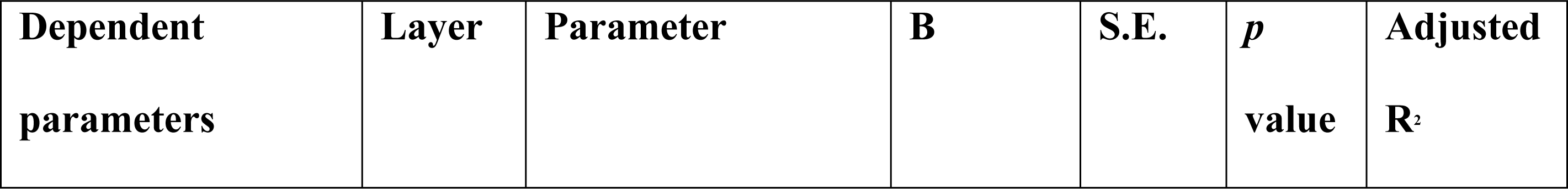
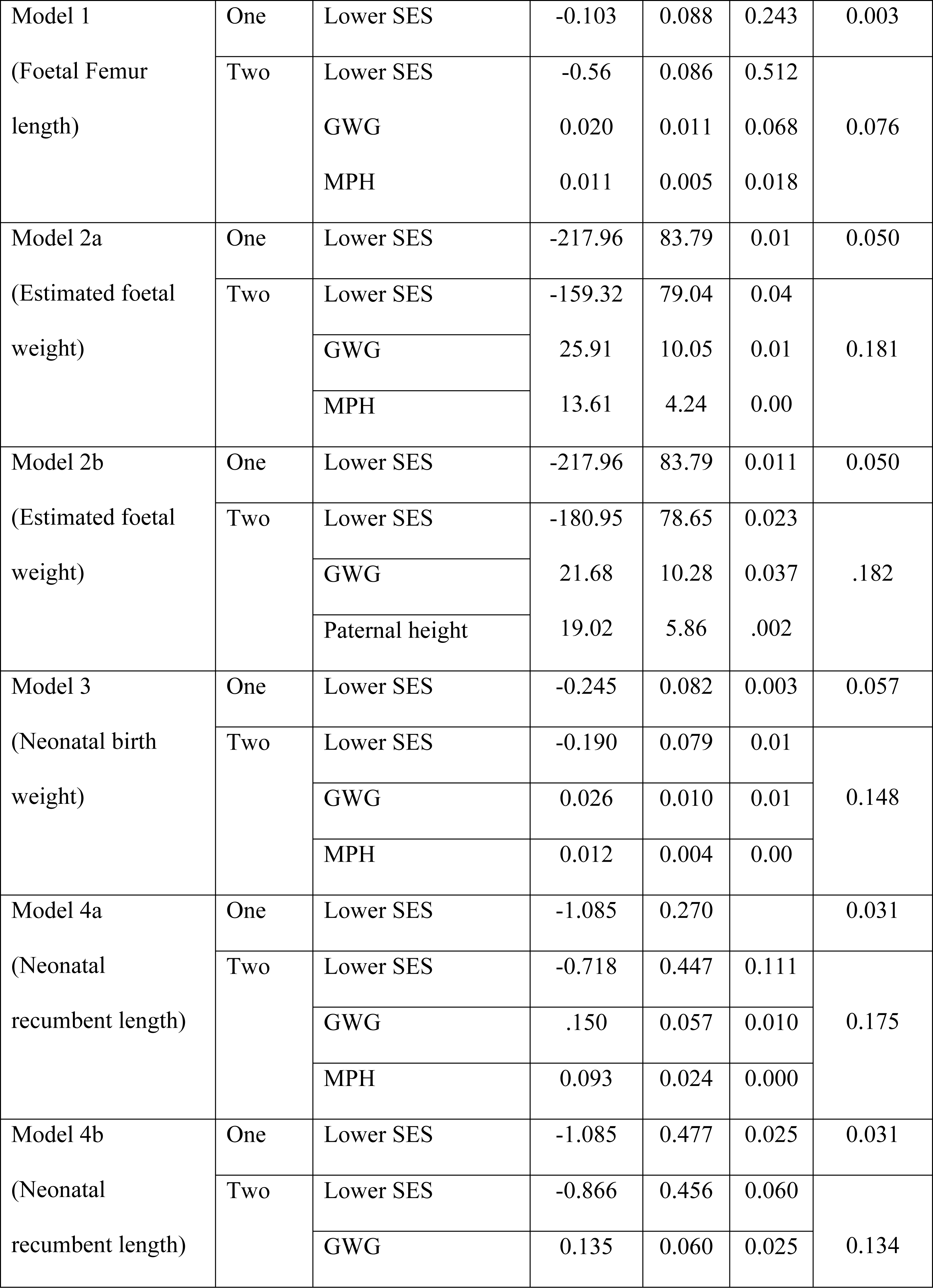
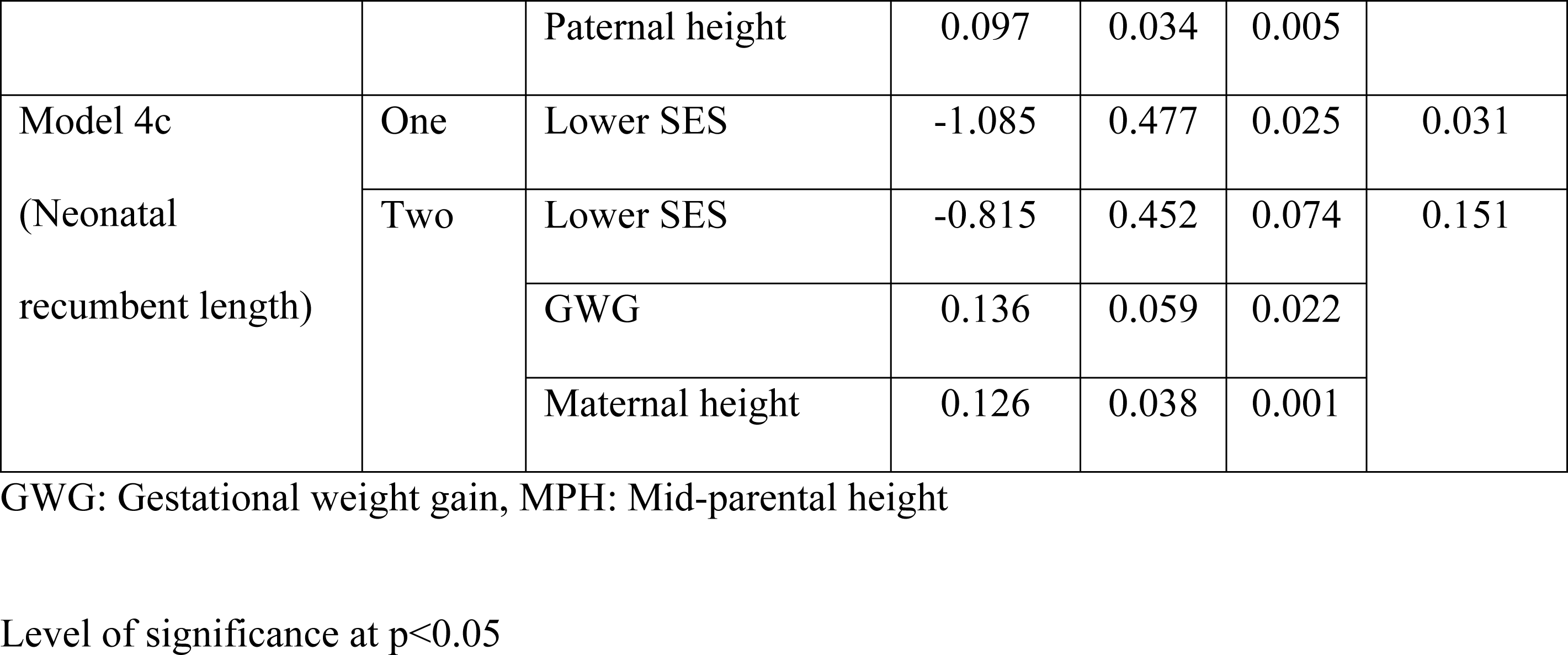
Determinants of foetal-neonatal growth parameters from MAI cohort

## Discussion

Parental influence on child growth has been a longstanding subject of interest in the field of maternal-new born and child health. This is one of the few studies conducted that highlights the importance of not only maternal, but also paternal anthropometry on foetal and neonatal growth. Our study has established gender-dependent relationships with parental anthropometry and neonatal growth, and further emphasized the influence of maternal gestational gains on foetal and neonatal growth.

Our study corroborates findings from multiple studies that highlight the influence of maternal weight gain on a neonate’s birth weight(8,23,24). Maternal nutritional status was found to be an important factor for determining neonatal growth in several studies however, data on findings from a socially disadvantaged population are scare. Zilko et al from a developed country setting found that the odds of an small-for-gestational age neonate decreased with improvement in gestation weight gain(23). Occurrence of adverse outcomes (small-for-gestational age, preterm birth, large-for-gestational age) was noted at the extreme ends of the spectrum (underweight and obese BMI mothers who did not gain adequate weight). Our study analysed the difference in foetal and neonatal growth between socially disadvantaged mothers who gained adequate and inadequate gestational weight. Mothers belonging to the inadequate gestational weight gain category had a higher risk of having neonates with low birth weight. Similar findings were reported by Perumal *et al,* wherein they reported that mothers who gained inadequate weight gave birth to low birth weight neonates and small for gestational age neonates(25).

Consistent with previous literature(26,27), we also observed a significant correlation between maternal pre-pregnancy BMI with foetal femur length from 20 weeks of gestation, unlike other foetal parameters which were significant since the 28 weeks of gestation. A study conducted in Vienna revealed that maternal height and BMI both strongly correlated with foetal size and neonatal size at birth(28). The associations were prominent from the first trimester onwards which was also observed in our study. These results highlight the influence of maternal nutritional characteristics on the length of the child since the foetal period. In the current study based in global south, an association was noted between maternal height and birth length, and birth weight and our results are thus in line with the narrative that tall mothers give birth to taller and heavier neonates, a result confirmed by Patra *et al* in a similar setting (29).

Our study highlighted a strong influence of maternal height on neonatal anthropometry, we also found that underweight mothers or those with shorter stature delivered low birth weight neonates as compared to their counterparts. A study by Witter *et al* and Britto *et* al corroborated these findings too, thus explaining the influence of maternal height on a neonate’s weight and not just length(30). It is generally hypothesized that maternal height is a strong predictor of gestational age and thereby neonatal outcomes viz. foetal weight and length at birth i.e., shorter mothers give birth early to neonates who have relatively shorter length and lower birth weight. Zhang *et al* aimed to study this hypothesis and they concluded that maternal height and foetal growth measures are in fact significantly associated, however, they attributed the association to foetal genetics(31).

In our study, we found that maternal family socio-economic status influences gestational weight and foetal-neonatal anthropometry and that the relationship between socio-economic status and these outcomes may be mediated by factors such as access to healthcare, dietary quality, and physical activity levels, etc. A few studies from different parts of the world highlight a similar phenomenon (32–34). We also observed that women with lower socio-economic status gave birth to neonates with lower birth weight than women with higher socio-economic status and Papazian *et al.* in their study highlighted this key finding(35). They also reported that maternal height and weight were important predictors of gestational weight gain, and that maternal education was associated with neonatal length and head circumference, which is yet another corroborating factor between their and our study. Another study conducted in India by Motwani *et al*. also reported similar results suggesting that maternal education was associated with gestational weight gain and neonatal anthropometry, and that the effect of maternal education on neonatal anthropometry was partially mediated by gestational weight gain, highlighting how socio-economic status is a strong determinant of foetal-neonatal growth. Data driven finding is chiefly highlighted in our study that women with lower socio-economic status are at higher risk of inadequate gestational weight gain, thus leading to a foetus of smaller size and consequently, it is hypothesized that they possibly give birth to neonates with lower birth weight.

Unlike the impact of maternal height and weight gains on foetal-neonatal growth, data on impact of paternal height on foetal-neonatal growth are scarce. Takagi *et al* reported the influence of paternal height on birth weight of the neonate wherein a gender dependant relationship was observed between paternal height and birth weight of the female neonate(36). An Indian study conducted with the aim of assessing the relationship between paternal and maternal height on foetal growth by Wills *et al*(2) concluded that paternal height was significantly correlated with foetal head circumference and femur length from 17 weeks of gestation onwards. In our study, paternal height significantly correlated with foetal femur length, abdominal circumference, head circumference, bi-parietal diameter and foetal weight from 28 weeks of gestation onwards where the correlation was much stronger at 36 weeks of gestation. Additionally, correlations between paternal height and neonatal birth weight and recumbent length were also found. We also observed that paternal height determined birth weight and recumbent length of the neonates unlike Morrison *et al* who concluded that father’s height significantly impacted only birth weight of the neonate(37). They found that up to 152 g variance in the neonatal weight can be attributed to paternal height with a larger effect when the mother was taller. However, in the current study we found that paternal height, independent of the maternal height influenced neonatal weight. Nahum *et al* corroborated findings of paternal height being an important factor in determining birth weight of the neonate. They concluded that only 250g of variance in birth weight can be explained by paternal height independently(38). They also found that with every 1cm increase in paternal height, birth weight increased by 10g. In the current study, an independent significant association was observed between paternal height and birth weight and length of the neonate. Similar findings have been reported by To *et al* and they concluded that paternal height could be a strong determinant of neonatal birth weight(39).

Our study demonstrated a relationship between fathers’ height and female offspring’s length and mother’s height and male neonate’s length, indicating a gender-dependent relationship. We cannot exclude the possibility of epigenetic factors, as a previous study in Japan indicated that paternal height may have a gender-dependent impact on the growth of the neonate(36). A study was conducted in a Norwegian population where they evaluated the association between parental height and weight with child’s height and weight at birth and until 2 years of age(40). They found that maternal weight significantly influenced femur length at 20 and 30 weeks of gestation whereas the current study found that maternal weight showed significant association from 28 weeks of gestation onwards. They also found a gender dependent relationship between parental height and neonate’s length at birth where paternal height influenced the female neonate’s length and maternal height influenced the male neonate’s length. Future studies can address the use of MPH for assessing combined effect of parental anthropometry on foetal and neonatal growth in different genders for a more comprehensive analysis.

We found that the mean gestational weight gain in our study participants was 9.9±3.7 kg. In a study by Zilko *et al,* gestational weight gain was around 14.2±6.9kg(23). The discrepancy noted in the GWG could be, as in the cohort studied by Zilko et al, a fairly large number of mothers belonged to a normal pre-pregnancy BMI category and very few women were underweight (in the present study 36.6% of women were underweight). Also, the mothers in that cohort complied with IOM weight gain guidelines leading to a greater GWG compared to ours where mothers largely belonged to an underweight pre-pregnancy BMI category and did not gain adequate weight during pregnancy. They also report a relationship between maternal gestational gains and neonatal size at birth, as also evidenced by our study.

To the best of our knowledge, this is one of the few studies being conducted in the global south among socially disadvantaged rural Indian population (low-middle income setting) in a longitudinal study design to assess the effects of parental anthropometry on intrauterine growth and neonatal outcomes. We have taken into account paternal as well as follow-up data on maternal nutritional status (thus considering maternal acute and chronic nutritional status), accounting for gestation-dependent growth of tissues. We have also assessed the influence of short maternal and paternal stature, and inadequate gestational weight gain on the growth of the child from rural settings which has not been well documented in rural India enough. One of the cornerstones of the current study is the gender-dependent relationship of parental nutritional status and growth of the neonate, which has been documented in Caucasian populations, but to date, data on Indian populations have been scarce.

Our study is not without limitations. A larger sample size would have made our model more generalizable, however, the power of our study was good enough. For calculating pre-pregnancy BMI, we relied upon participant recall for weight before pregnancy, which could have generated recall bias; however, we cross-checked weight in the medical record books as well as measured actual first trimester weight to omit potential discrepancies. Moreover, sonographies were conducted by multiple ultrasonologists, given the vast distances between data collection centres and participants’ residences which could have resulted in variations in readings; however, the sonographies were eventually examined by one examiner to nullify potential error-based variation. Due to the unavailability of paternal weight, associations between acute paternal nutritional status and growth of the child could not be made. Future recommendation should highlight on conducting longitudinal prospective cohort studies with a larger sample size whilst examining the impact of parental birth anthropometry and socio-economic status on the growth of their child in-utero and at birth, this should help researchers identify and understand the phenomenon of inter-generational malnutrition which would in-turn reduce the incidence of adverse parental-foetal-neonatal growth outcomes.

## Conclusion

Our study reveals the impact of parental socioeconomic status, anthropometry as well as maternal pregnancy weight gain on foetal-neonatal weight and length. Given that the majority of mothers who were underweight during pre-pregnancy had inadequate gestational weight gain, this study underlines the need for interventions pre-pregnancy and during pregnancy to ensure optimal weight gain as this has long-term implications for foetal and neonatal outcomes. In addition, interventions aimed at improving socioeconomic status and maternal education, in particular, may also contribute to improvements in gestational weight gain and foetal-neonatal outcomes.

## Data Availability

Data that supports the findings are available from the HCJMRI, pending approval from ethics committee.

## Acknowledgements and disclosures

We are grateful to all our study participants for their active and voluntary participation, ultrasonologists and primary health care centre heads of study centres for their support and encouragement. Mugdha Deshpande was funded by a fellowship grant from University Grants Commission, Government of India. NTA Ref. no.: 220510107214 (NET JRF Dec 2021 and June 2022).

## Author contributions

Mugdha Deshpande: Conceptualization, Methodology, Software, Data curation, Writing- Original draft preparation, Visualization, Investigation

Demi Miriam: Conceptualization, Writing- Original draft preparation, Reviewing and Editing

Nikhil Shah: Conceptualization, Writing- Original draft preparation, Visualization, Investigation, Validation, Reviewing and Editing

Neha Kajale: Writing- Original draft preparation, Supervision, Validation, Reviewing and Editing

Jyotsna Angom: Supervision, Validation, Reviewing and Editing

Jasmin Bhawra: Investigation, Validation, Writing- Reviewing and Editing

Ketan Gondhalekar: Investigation, Validation, Data curation, Software

Anuradha Khadilkar: Conceptualization, Methodology, Writing- Original draft preparation, Visualization, Investigation, Supervision, Validation, Reviewing and Editing

Tarun Katapally: Writing- Investigation, Validation, Supervision, Reviewing and Editing

## Supporting information

Ethics approval letter has been obtained from the institutional ethics committee which has been uploaded on the editorial manager.

